# The impact of preoperative imaging strategies in EGFR-mutant non-small cell lung cancer: a multicenter retrospective review

**DOI:** 10.1101/2025.10.30.25339077

**Authors:** Jonathan W. Lee, Michael Rafizadeh, Stephanie Bogdan, Nasser Altorki, Christine Garcia, Ashish Saxena, Bobak Parang

**Affiliations:** Division of Hematology and Oncology, Department of Medicine, Weill Cornell Medical College, New York, NY 10021; Department of Cardiothoracic Surgery, Weill Cornell Medical College, New York, NY 10021; Division of Medical Oncology, Department of Internal Medicine, The Ohio State University Comprehensive Cancer Center, Columbus, OH, 43210

**Keywords:** EGFR, non-small cell lung cancer, preoperative imaging, ADAURA

## Abstract

**Background:** The ADAURA trial demonstrated an overall survival benefit with adjuvant osimertinib in epidermal growth factor receptor-mutant (EGFR-mut) non-small cell lung cancer (NSCLC). As only 50% of patients underwent a preoperative brain MRI scan and PET-CT rates were not reported, concerns were raised that suboptimal staging could have led to the inclusion of patients with metastatic disease. There is, however, limited data on the real-world rates and impact of preoperative PET-CT and brain MRIs in patients with EGFR-mut NSCLC. We sought to characterize preoperative imaging rates and their impact on staging.

**Patients and Methods:** We retrospectively analyzed patients with EGFR-mut NSCLC who were evaluated for surgery at 3 academic New York hospitals from 2016-2021.

**Results:** Between 2016 and 2021,109 patients with EGFR-mut NSCLC underwent preoperative evaluation. Of these patients, 107 underwent a PET-CT and 15 (14%) were found to have stage IV disease. Sites of metastatic disease were bone (11), pleura (2), liver (1), and intrapulmonary (1). Thirty-nine patients had stage II or III disease; of those, 23 underwent a brain MRI, which revealed brain metastases in 3 patients. In total, the use of PET-CT and brain MRIs for patients evaluated for surgery upstaged 17% (18/109) of patients to metastatic disease and resulted in 83% (90/109) of patients undergoing surgery.

**Conclusions:** The use of preoperative PET-CT and brain MRIs led to the detection of metastatic disease in 17% of patients with EGFR-mut NSCLC. These findings underscore the critical role of comprehensive preoperative imaging for clinical practice and clinical trials.

## Introduction

The international, randomized phase 3 ADAURA trial demonstrated adjuvant osimertinib improved overall survival (OS) in patients with resected stage IB to IIIA epidermal growth factor receptor-mutated (EGFR-mut) non-small cell lung cancer (NSCLC).^1,2^ The trial did not mandate preoperative positron-emission tomography/computed tomography (PET-CT) scans or brain magnetic resonance imaging (MRI), raising concerns that patients were suboptimally staged.^3,4^ The National Comprehensive Cancer Network (NCCN) has recommended PET-CT for NSCLC since 2006 since PET-CT has shown superiority to CT alone in detecting metastatic disease.^5–8^ Similarly, since 2009, the NCCN has recommended brain MRIs for patients with stage II-IV NSCLC because of its improved sensitivity compared to CT.^9–12^

Fifty-one percent of patients in the ADUARA trial underwent a brain MRI; the rates of PET-CT imaging have not been disclosed.^13^ Criticisms of the trial have argued that suboptimal staging would lead to the inclusion of patients with metastatic disease and favor the investigational arm,^3,4^ diminishing the applicability of the results to clinicians in the United States (US) given NCCN guidelines. However, there is limited data on real-world rates of PET-CT and brain MRI imaging for patients with EGFR-mut NSCLC who are evaluated for surgical resection, and the practice patterns in the US are not clear. Moreover, there is little reported data on the *impact* of staging with preoperative PET-CT and brain MRI for patients with EGFR-mut NSCLC. Prior reports demonstrating the superiority of PET-CT and brain MRI were mostly conducted in the early 2000s using older imaging technology and prior to routine genomic sequencing of NSCLC.^14–16^

Given the number of ongoing trials focused on using targeted therapies in the adjuvant setting, it is important to establish the clinical practices of preoperative staging for patients with targetable mutations and the effect of preoperative PET-CT and brain MRIs on upstaging patients with early-stage NSCLC.

Here, we report rates of preoperative PET-CT and brain MRI and their impact on upstaging patients with EGFR-mut NSCLC who were evaluated for surgery across 3 US medical centers.

## Methods

### Study Design and Patients

This retrospective study evaluated patients diagnosed with EGFR-mut NSCLC between 2016 and 2021 at 3 academic New York hospitals: New York-Presbyterian (NYP) Weill Cornell, NYP-Queens, and NYP-Brooklyn Methodist.

To be included in the study, patients: (1) were histologically and/or cytologically confirmed to have NSCLC; (2) had an activating EGFR mutation that was either an exon 19 deletion or an exon 21 L858R point mutation; and (3) were evaluated for curative-intent surgery. Patients with evidence of metastatic disease based on initial clinical presentation or initial non-PET-CT imaging were excluded from this study.

## Results

### Patient and Disease Characteristics

A total of 109 patients were included for final analysis (**Table 1**). Median age at diagnosis was 70 years (range 43-89), and 81 patients (74%) were female. Forty-seven patients (43%) were of Asian ethnicity and 63 (58%) had no history of smoking. Eastern Cooperative Oncology Group (ECOG) performance status was between 0-1 for 88 patients (81%) at diagnosis. Forty-four patients (40%) had a TP53 co-mutation.

**Table 1.** Patient Characteristics.

|  |  |
| --- | --- |
| **Evaluable patients, *N*** | 109 |
| **Median age (range), years** | 70 (43-89) |
| **Female, *n* (%)** | 81 (74) |
| **ECOG 0-1, *n* (%)** | 88 (81) |
| **Never smoking history, *n* (%)** | 63 (58) |
| **Ethnicity, *n* (%)** |  |
| Asian | 47 (43) |
| White | 46 (42) |
| Unknown | 7 (6) |
| Hispanic | 6 (6) |
| Black | 3 (3) |

**Abbreviations:** ECOG – Eastern Cooperative Oncology Group

### PET-CT Rates and Impact on Preoperative Staging

Initial clinical staging of lung cancer was done with non-contrast CT scans for 66 patients (61%), while 30 patients (28%) had a CT scan with contrast (**Table 2**). The majority of patients had stage I disease (**Table 2**).

**Table 2.** Initial Imaging and Stage.

|  |  |
| --- | --- |
| **Total patients, *N*** | 109 |
| **Initial imaging, *n* (%)** |  |
| Non-contrast CT | 66 (61) |
| Contrast CT | 30 (28) |
| MRI | 1 (1) |
| **AJCC stage, *n* (%)** |  |
| IA | 47 (43) |
| IB | 23 (21) |
| IIA | 11 (10) |
| IIB | 5 (5) |
| IIIA | 15 (14) |
| IIIB | 4 (4) |
| IIIC | 1 (1) |
| IVA | 1 (1) |

**Abbreviations:** CT – computed tomography; AJCC – American Joint Committee on Cancer

Of the 109 patients, 107 (98%) had a PET-CT as part of their surgical evaluation. Of the 107 patients who had a PET-CT, a total of 30 (29%) had a change in their clinical staging and were “upstaged.” Fifteen patients (14%) were upstaged from stage I and II disease because of new thoracic lymphadenopathy or an increase in the primary lesion size (**Table 3**), and another 15 patients (14%) were upstaged to stage IV.

**Table 3.** Effects of Preoperative PET-CT on Staging.

|  |  |
| --- | --- |
| **Evaluable Patients, *N*** | 109 |
| **Obtained preoperative PET-CT, *n* (%)** | 107 (98) |
| **Change in clinical stage due to PET-CT (non-stage IV), *n* (%)** | **15 (14)** |
| Due to change in size of primary lesion | 4 (27) |
| Due to new thoracic lymphadenopathy | 11 (73) |
| **Upstaged to stage IV by PET-CT, n (%)** | **15 (14)** |
| Sites of metastatic disease |  |
| Bone | 11 (73) |
| Pleural disease | 2 (13) |
| Liver | 1 (7) |
| Intrapulmonary disease | 1 (7) |

**Abbreviations:** PET-CT – positron-emission tomography/computed tomography

Among these 15 patients who were diagnosed with metastatic disease, 7 were initially diagnosed as stage I by CT imaging alone. Metastatic sites of disease that PET-CT identified were bone (11), pleura (2), liver (1), and intrapulmonary (1) (**Figure 1**).

**Figure 1.** Sites of metastatic disease identified by PET-CT

### Impact of Brain MRI on Preoperative Staging

The NCCN guidelines recommend brain MRI for all patients with NSCLC except for patients with stage I disease. Among our patient cohort, there were 39 patients that fit criteria for the NCCN recommendation of preoperative brain MRI (stage II and III) (**Table 4**). Twenty-three of those patients (59%) underwent a brain MRI, which revealed central nervous system (CNS) metastases in 3 patients (7%). These 3 patients did not have metastatic disease detected by PET-CT. Among the 70 clinical stage I patients, 16 underwent a brain MRI, which identified CNS metastases in 2 patients.

**Table 4.** Effects of Preoperative Brain MRI.

|  |  |
| --- | --- |
| Evaluable patients, N | 109 |
| Recommended preoperative brain MRI per NCCN guidelines | 39 |
| Obtained preoperative brain MRI, n (%) | 23 (59) |
| Found to have CNS disease, n (%) | 3 (7) |

**Abbreviations:** MRI – magnetic resonance imaging; NCCN – National Comprehensive Cancer Network; CNS – central nervous system

In total, the use of both PET-CT and brain MRI for patients evaluated for surgery upstaged 18 out of 109 patients (17%) to metastatic disease (**Figure 2**). In our patient cohort, 90 patients (83%) ultimately underwent curative-intent surgery.

**Figure 2.** (A) Percentage of patients upstaged by PET-CT or brain MRI to stage IV. (B) Initial stage based on CT-only of patients who were ultimately diagnosed with stage IV disease by PET-CT or brain MRI

## Discussion

As treatment strategies for early-stage NSCLC continue to evolve and targeted therapies are tested in the adjuvant setting,^13,17^ it is important to define the impact of preoperative imaging modalities in order to assess clinical trials. Considering the ADAURA trial and its criticisms, we sought to determine our institution’s preoperative imaging practices for patients with EGFR-mut NSCLC and how they relate to ADAURA.

Our study shows that 98% of our cohort underwent a PET-CT prior to surgery between 2016-2021. The rates of patients who underwent a preoperative PET-CT in ADAURA have not yet been published, although all patients were mandated to have a postoperative chest, abdomen, and pelvis CT prior to adjuvant treatment. Strikingly, in our cohort, 14% of patients who underwent PET-CT imaging as part of a workup for curative-intent surgery were diagnosed with metastatic disease that was not detected on initial CT imaging. Interestingly, in 73% of the patients who were diagnosed with stage IV disease, the site of metastases was bone. Given PET-CT imaging is superior to CT-alone imaging for detecting bone metastases in NSCLC,^18^ our data suggests that patients with occult bone metastases were likely included in the ADAURA trial. Another 20% of patients in our cohort were diagnosed with metastatic disease due to either pleural or intrapulmonary metastases that were not identified on initial CT scans. Almost half of all patients who were diagnosed with stage IV disease were initially stage I based on CT imaging, underscoring the importance of PET-CT for patients with EGFR-mutant NSCLC.

In the ADAURA trial, 51% of all patients underwent a brain MRI. At our institution, of the patients for whom the NCCN recommends a brain MRI (stage II and stage III), 59% had a preoperative brain MRI. Brain MRI revealed 7% of patients had CNS disease. Importantly, these patients did not have metastases detected by PET-CT and were upstaged solely by brain imaging. We were surprised by the relatively low rates of preoperative brain MRIs in our patient cohort. One patient declined a brain MRI because of claustrophobia. What accounts for the others is not clear. Routine brain MRIs in asymptomatic surgical patients is controversial. Although the NCCN recommends it in patients with stage II and III disease, not all professional societies agree.^19,20^ In our study, we assessed clinical practices between 2016-2021 to compare them to practices during the enrollment of patients in the ADAURA trial, which enrolled patients between 2015 and 2019. Routine next-generation sequencing (NGS) was not necessarily performed for patients with resectable disease during this period, which may have influenced whether patients were considered higher risk for brain metastases (i.e., harbored an EGFR-mutation or ALK-fusion) and required preoperative brain MRIs.

Our study has several limitations. First, we focused on patients at academic centers, which does not necessarily reflect broader practices across the US. Second, this was a retrospective study that relied on chart review, carrying inherent biases. Third, this study was conducted intentionally on patients who were evaluated between 2016-2021 to evaluate practice strategies in parallel to ADAURA patient enrollment. Many societal guidelines now recommend preoperative NGS on lung tumors to identify oncogenic drivers, which may increase the frequency of preoperative brain MRIs given the higher rate of brain metastases in patients with EGFR-mut NSCLC.

Our retrospective study showed that almost all patients who underwent surgical evaluation for EGFR-mutant NSCLC underwent PET-CT imaging while over half obtained a preoperative brain MRI. Preoperative PET-CT and brain MRI identified metastatic disease in 17% of all patients, emphasizing the importance of mandating preoperative PET-CT and brain MRIs in clinical trials for resectable NSCLC.

### Clinical Practice Points

- PET-CT identified metastatic disease in 14% of patients with EGFR-mutant NSCLC who underwent evaluation for curative resection.
- Disease sites were primarily bone, which is challenging to detect by CT imaging.
- Brain MRI imaging was performed in 23/39 patients for whom the NCCN recommends imaging.
- Brain MRI detected brain lesions in 7% of patients with EGFR-mutant NSCLC who underwent evaluation for surgery.
- Patients with EGFR-mutant NSCLC are at high risk for occult metastatic disease and should undergo preoperative PET-CT and brain MRI imaging.

## Data Availability

All data produced in the present work are contained in the manuscript

## Acknowledgments

The authors would like to thank Angela Dahlberg, editor in the OSUCCC Division of Medical Oncology, for editing this manuscript.

## Author Contributions

**Jonathan W. Lee:** Writing—original draft, Data curation, Formal analysis, methodology **Michael Rafizadeh**: Data curation, **Stephanie Bogdan:** Data curation, **Nasser Altorki**: Writing-review and editing, **Christine Garcia**: Writing-review and editing **Ashish Saxena**: Writing-review and editing, **Bobak Parang**: Writing-original draft, Writing-review and editing, Investigation, Conceptualization, Formal analysis

